# A New Saliva-Based Lateral-Flow SARS-CoV-2 IgG Antibody Test for mRNA Vaccination

**DOI:** 10.1101/2021.06.11.21258769

**Authors:** Dingying Shan, Jessica Hsiung, Kevin P. Bliden, Su Zhao, Tao Liao, Guoxing Wang, Shuanglin Tan, Tiancheng Liu, Deepika Sreedhar, Jessica Kost, Shin Ting Chang, Wei Po Yuan, Udaya Tantry, Paul Gurbel, Meijie Tang, Hongjie Dai

## Abstract

Sensitive detection of IgG antibodies against SARS-CoV-2 is important to assessing immune responses to viral infection or vaccination and immunity duration. Antibody assays using non-invasive body fluids such as saliva could facilitate mass testing including young children, elderly and those who resist blood draws, and easily allowing longitudinal testing/monitoring of antibodies over time. Here, we developed a new lateral flow (nLF) assay that sensitively detects SARS-CoV-2 IgG antibodies in the saliva samples of vaccinated individuals and previous COVID-19 patients. The 25-minute nLF assay detected anti-spike protein (anti-S1) IgG in saliva samples with 100% specificity and high sensitivity from both vaccinated (99.51% for samples ≥ 19 days post 1st Pfizer/BioNTech or Moderna mRNA vaccine dose) and infected individuals. Antibodies against nucleocapsid protein (anti-NCP) was detected only in the saliva samples of COVID-19 patients and not in vaccinated samples, allowing facile differentiation of vaccination from infection. SARS-CoV-2 anti-S1 IgG antibody in saliva measured by nLF demonstrated similar evolution trends post vaccination to that in matching dried blood spot (DBS) samples measured by a quantitative pGOLD lab-test, enabling the nLF to be a valid tool for non-invasive personalized monitoring of SARS-CoV-2 antibody persistence. The new salivary rapid test platform can be applied for non-invasive detection of antibodies against infection and vaccination in a wide range of diseases.

## Introduction

The COVID-19 pandemic by the severe acute respiratory syndrome coronavirus 2 (SARS-CoV-2) has been presenting a major global health crisis^1-6^. It is clear that mass vaccination offers a viable path back to normalcy^7,8^. As of May 2021, 17 different COVID-19 vaccines across four platforms are rolled out across the globe, with more than 300 additional vaccine candidates still in development. Over 1.8 billion vaccine doses have been administered worldwide, raising the hope of emerging from the pandemic^9^. However, questions linger on the duration of the protection offered by vaccination against COVID-19. Since antibody response and persistence vary among individuals and between the different vaccines in circulation, antibody assessment at the individual level could help to evaluate antibody persistence and the necessity of booster doses. Various antibody tests have been developed using enzyme-linked immunosorbent assay (ELISA)^10^, nanostructured plasmonic gold (pGOLD) assay^11^, chemiluminescence (CLIA)^12^ and lateral flow (LF) technologies^13^. Antibodies against spike proteins have shown high correlation with neutralizing antibodies for COVID-19, offering a simple, first-line assessment of immunity^14-22^. Currently, most SARS-CoV-2 antibody tests require professional operations and invasive blood/serum processes, limiting the accessibility and frequency of testing. To facilitate personalized monitoring of SARS-CoV-2 antibody persistence, non-invasive and easy testing strategies are desired. Saliva represents an appealing biofluid for non-invasive SARS-CoV-2 antibody detection, with the ease of self-collection and is pain-free. Previous work has shown that IgG antibodies in saliva correlated well with those in blood for various infectious diseases including SARS-CoV-2^11,23-32^. However, a challenge of saliva sample testing is the much lower concentration of antibodies present than in serum, plasma and blood, demanding high analytical sensitivity of detection methods^33^. The high viscosity of saliva samples also presents a challenge to flow-based rapid testing. At this time, no saliva-based antibody LF assay exist for SARS-CoV-2 vaccination or infection assessment at the point-of-care (POC).

Here, we developed a new lateral flow (nLF) test that is highly sensitive for SARS-CoV-2 IgG antibody detection in saliva. In conventional antibody LF assays, when a biofluid sample containing targeting antibodies is added, the sample flows through a conjugate pad containing gold (Au)-antigen complexes to form Au-antigen-antibody complexes, which migrate towards the detection zone and bind to the test line containing immobilized anti-human antibodies. A readable test line (whether by visual or reader-based detection) appears due to the complex-bound Au nanoparticles suggests a positive test result^34^. A limitation to this LF approach is that biofluid samples always contain abundant non-specific human antibodies that compete with the specific antibodies captured by the Au-antigen complex for the anti-human antibody binding sites on the test line, causing low analytical sensitivity and non-linearity of the assay. Moreover, saliva samples are viscous, requiring high degrees of dilution in order to run on conventional LF strips, further lowering the analytical sensitivity of saliva-based LF tests.

For our nLF approach, we first developed and optimized Au nanoparticle-antigen (or magnetic Fe3O4 core/Au shell nanoparticle-antigen) complexes, then incubated with a saliva sample in a reaction tube to capture antibodies specific to the antigen, removed the non-specific antibodies (by micro-centrifuging or magnetic purification), and then applied the Au-antigen-antibody complexes to a LF test card containing anti-human IgG signal line for detection. The entire assay time was about 25 minutes. A portable, economic micro-centrifuge or magnetic rack was employed to remove the non-specific antibodies, making the assay user-friendly in POC settings. The nLF assay affords high anti-SARS-CoV-2 S1 IgG sensitivity (> 99%) with vaccinated saliva samples collected ≥ 19 days post the first Pfizer/BioNTech or Moderna mRNA vaccine dose accompanied by 100% specificity. By applying both SARS-CoV-2 spike protein S1 subunit and nucleocapsid (also known as nucleoprotein, NCP) to make Au-antigen complexes (Au-S1 and Au-NCP) for nLF assays, we differentiated saliva samples of SARS-CoV-2 vaccinated subjects from COVID-19 infected subjects. Importantly, anti-S1 antibodies in saliva showed a similar trend as those measured by a quantitative serology test performed on the pGOLD lab-test platform ^11^. Overall, the rapid nLF assay platform can be easily implemented at various POC sites including clinical laboratories, pharmacies, or eventually at home for non-invasive, highly accurate SARS-CoV-2 IgG monitoring.

## Results

### A new lateral flow (nLF) assay platform

The concentration of IgG antibodies in human saliva is much lower (∼15 µg mL^-1^) than in serum or blood (∼15 mg mL^-1^), so antibodies against SARS-CoV-2 spike protein in saliva are more difficult to detect than in sera of COVID-19 infected individuals^11,24,27,33^. Several lab-based assays have been developed to meet this challenge, but not for saliva-based POC LF tests.

To develop our nLF SARS-CoV-2 IgG antibody saliva test, we first prepared and optimized Au nanoparticle-antigen complexes. The Au nanoparticles were either pure gold (∼ 50 nm) or magnetic gold comprised an iron-oxide Fe3O4 core and an Au shell (see Methods). When the Au-antigen (antigen = S1 or NCP of SARS-CoV-2) complexes were incubated with positive saliva samples, IgG antibodies specific to the viral antigen were captured to form Au-antigen-IgG complexes **(Figure 1a (i-iii))**. A centrifuging or magnetic purification step was applied afterwards to collect the Au-antigen-IgG complexes and remove the unbound, non-specific antibodies. The resulting Au-antigen-IgG complexes were dispersed in a running buffer and added to a specially designed LF testing card to flow through the signal and control lines **(Figure 1a (iv))**. The card was comprised of a sample pad, a conjugate pad with pre-deposited colloidal gold nanoparticle-biotinylated BSA (Au-biotin-BSA) complexes, a test line immobilized with anti-human IgG, a control line immobilized with streptavidin, and a terminal absorbent pad **(Figure 1b)**. When antigen-specific IgG positive samples were used for the nLF, both test and control lines were detectable on the card as red lines, indicating a positive test, while negative samples or buffer alone only produced a red line in the control line **(Figure 1c)**. The step of removing unbound non-specific antibodies by micro-centrifuging or magnetic purification was critical to preventing competing antibodies in the sample from binding to the test line, drastically enhancing the detection sensitivity of specific antibodies over the traditional LF assay. The total nLF assay time was ∼ 25 minutes.

**Figure 1.**
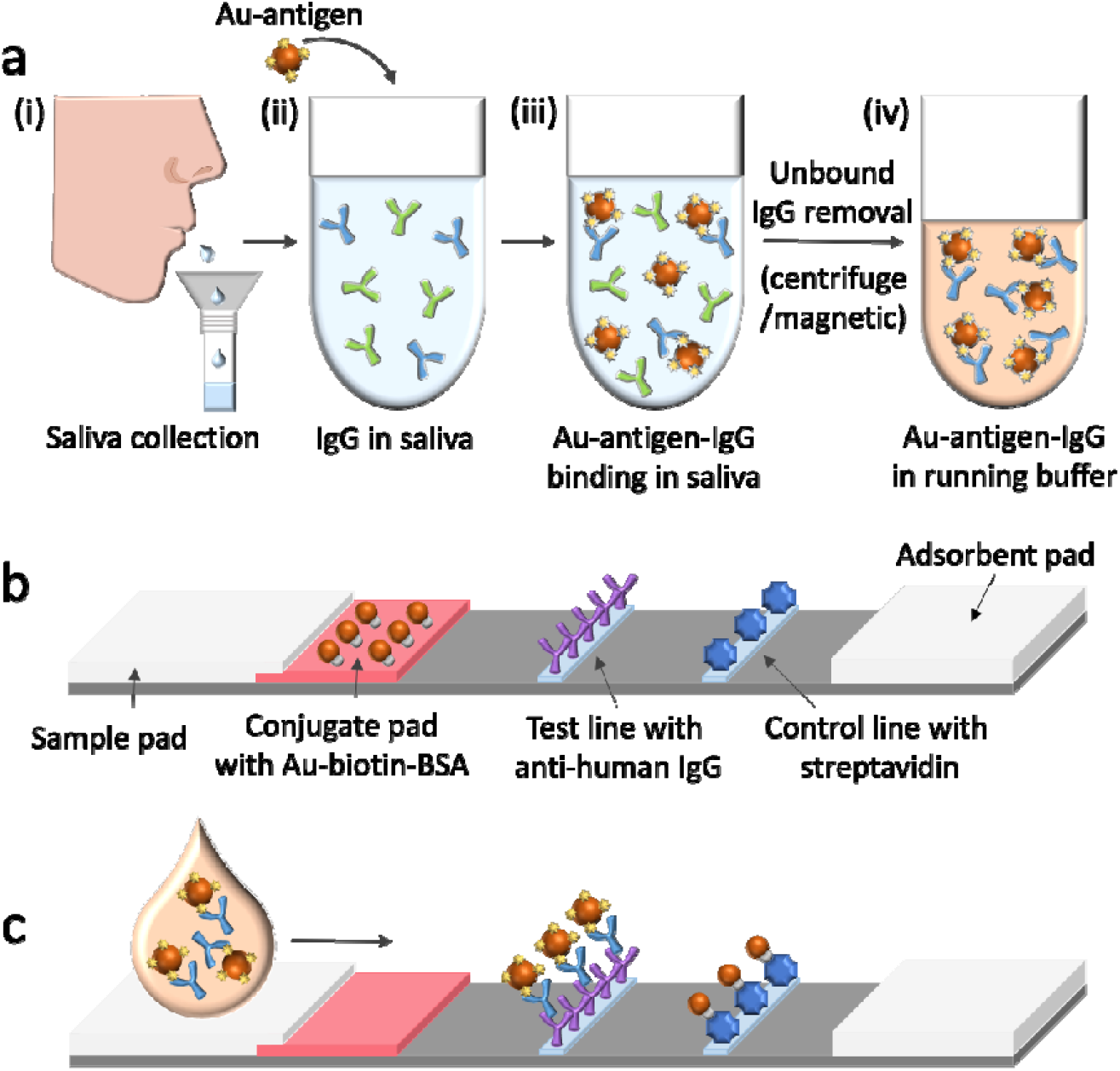
A new lateral flow (nLF) test for SARS-CoV-2 antibody testing in saliva. **a**, complex formation. After saliva sample collection (i), Au-antigen (antigen = S1 or NCP of SARS-CoV-2) complexes were incubated with the sample (ii) and capture antigen-specific IgG antibodies to form Au-antigen-IgG complexes (iii). A centrifuging or magnetic purification step was applied afterwards to collect the Au-antigen-IgG complexes and remove non-specific antibodies. The obtained Au-antigen-IgG complexes were dispersed in a PBS buffer before being added to a specially designed LF testing card. **b**, Design of the nLF testing card. The card is comprised of a sample pad, a conjugate pad with pre-deposited colloidal gold nanoparticle-biotinylated BSA (Au-biotin-BSA) complexes, a test line immobilized with anti-human IgG, a control line immobilized with streptavidin, and a terminal adsorbent pad. **c**, Operation of the nLF assay. When antigen-specific IgG positive samples are used for the nLF, both test and control lines are detectable on the card, indicating a positive test, while negative samples or buffer alone only produced a red line in the control line.

### Performance of the nLF assay

We first compared the detection limit of nLF to that of a conventional LF test with FDA Emergency Use Authorization (MidaSpot™ COVID-19 Antibody Combo Detection Kit, or MidaSpot, developed by Nirmidas Biotech Inc.). We spiked purified recombinant anti-SARS-CoV-2 spike protein IgG into a negative healthy serum sample at various concentrations to determin the limit of detections by nLF and MidaSpot. MidaSpot showed positive signals for 4X and lower dilutions **(Figure S1a)**. The nLF detected anti-S1 IgG even in the 256X diluted sample, affording a ∼ 64 times lower detection limit than the conventional LF approach **(Figure S1b)**, owing to the removal of excess, non-specific serum IgG by centrifugation or magnetic suction.

We also spiked purified recombinant anti-SARS-CoV-2 spike protein IgG into a negative healthy saliva sample and performed serial dilutions to establish the nLF assay detection limit for anti-S1 IgG in saliva. By quantifying the signal intensity on the test line (see Methods), we obtained an estimated antibody detection limit of ∼ 0.30 µg mL^-1^, ∼ 50-fold lower than the total antibody concentration (∼15 µg mL^-1^) in human saliva **(Figure 2a)**^33^.

**Figure 2.**
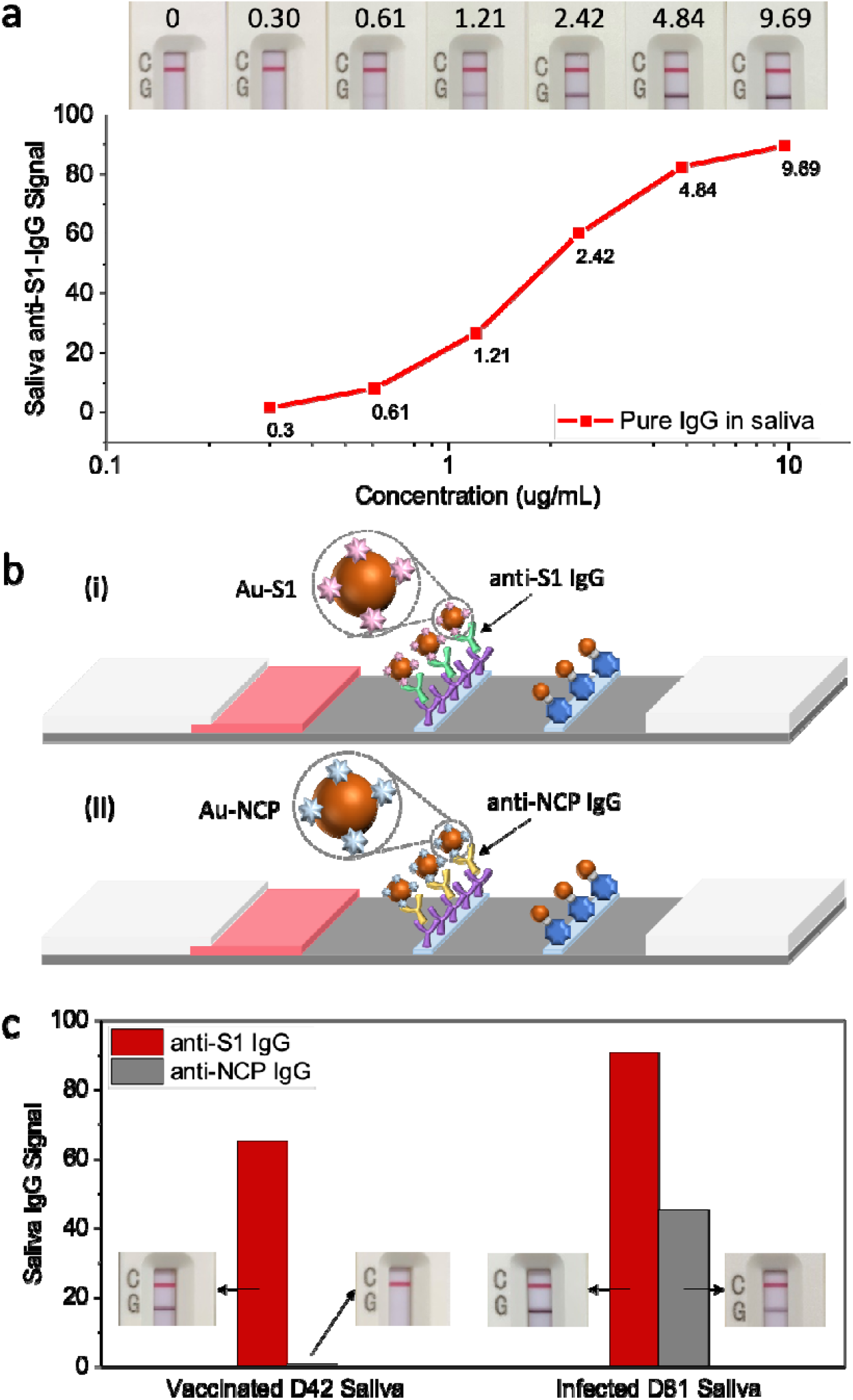
Highly sensitive and multifunctional nLF test for SARS-CoV-2 antibody testing in saliva. **a**, Detection limit of anti-SARS-CoV-2 spike protein IgG in saliva. Purified recombinant anti-SARS-CoV-2 spike protein IgG were spiked into a negative healthy saliva sample and serial dilutions were performed to establish the nLF assay detection limit for anti-S1 IgG in saliva. By quantifying the signal intensity on the test line (see Methods), we obtained an estimated antibody detection limit of ∼ 0.30 µg mL^-1^. **b**, Schematic representation of nLF for anti-S1 IgG detection (i) and anti-NCP IgG detection (ii). **c**, Representative testing results and IgG signals obtained from nLF for vaccinated and infected saliva samples. A typical vaccinated saliva sample (42 days post 1st RNA vaccine dose from Moderna) shows positive anti-S1 IgG and negative anti-NCP IgG, while an infected saliva sample (81 days post symptom onset) shows both positive anti-S1 IgG and positive anti-NCP IgG.

We next focused on nLF assays of SARS-CoV-2 antibodies in saliva samples from vaccinated and COVID-19 infected cohorts. The major SARS-CoV-2 antigenic targets of human IgG are the spike protein and nucleocapsid (NCP). The SARS-CoV-2 spike protein is used as the target antigen and encoded in the mRNA vaccines from Pfizer/BioNTech and Moderna. Individuals who have been successfully vaccinated by the one of the two mRNA vaccines, without prior COVID-19 infection, should only test positive for anti-S1 IgG but not for anti-NCP IgG ^35-37^. However, COVID-19 infected patients are known to test positive for both anti-S1 IgG and anti-NCP IgG ^38^. The nLF assay platform is flexible for testing human IgG antibodies against any protein target by simply changing Au-antigen complexes while using the same test card. Using Au-S1 and Au-NCP complexes, we readily developed nLF tests for evaluating IgG antibodies against SARS-CoV-2 spike protein and NCP in saliva samples **(Figure 2b)**. With these tests, we observed a typical vaccinated saliva sample (42 days post 1st RNA vaccine dose from Moderna) exhibiting positive anti-S1 IgG and negative anti-NCP IgG, while an infected saliva sample (81 days post symptom onset) showing both positive anti-S1 IgG and positive anti-NCP IgG **(Figure 2c)**. The same pattern was observed for larger numbers of vaccinated (20) and infected samples (20) tested.

### Salivary antibody patterns: vaccination vs. infection

To fully establish the nLF assay for saliva anti-S1 IgG testing, we tested 34 unvaccinated healthy saliva samples without prior COVID-19 infection, 20 saliva samples from PCR confirmed COVID-19 patients (16 samples were collected ≥ 15 days post symptom onset, 1 sample was collected 11 days post symptom onset, and 3 samples had no information regarding the number of days between symptom onset to sample collection), and 205 vaccinated (with mRNA vaccines from Pfizer/BioNTech and Moderna) saliva samples (collected ≥ 19 days post 1st vaccine shot from subjects without prior COVID-19 infection). For salivary SARS-CoV-2 anti-S1 IgG, a cut-off value of 0.921 T-line intensity unit was determined via the receiver operating characteristic (ROC) curve under the criteria of 100% specificity and 99.6% sensitivity **(Figure S2a)**. The nLF saliva assay afforded 100% specificity without false positive for the 34 unvaccinated COVID-19 free cohort. Salivary SARS-CoV-2 anti-S1 IgG antibodies were detected in 20 out of 20 COVID-19 infected subjects (100%), and in 204 out 205 vaccinated subjects (99.51%). For the 205 vaccinated subjects, only 1 saliva sample on day 21 post the 1st vaccine dose/prior to the 2nd dose was tested negative.

For the anti-NCP IgG nLF assay, a cut-off value was determined by testing saliva samples from 12 unvaccinated COVID-19 free individuals and 20 COVID-19 infected individuals. The ROC analysis gave a cutoff value of 1.004 intensity unit under the criteria of 100% specificity and 100% sensitivity **(Figure S2b)**. All of the 20 COVID-19 infected saliva samples were tested positive for both anti-S1 IgG (100%) and anti-NCP IgG (100%) **(Figure 3a)**. In contrast, of the 20 vaccinated saliva samples tested by nLF on day 42-55 post 1st vaccine dose, 0 out of 20 (0%) was tested positive for anti-NCP IgG **(Figure 3b)**, and all 20 (100%) were tested positive for anti-S1 IgG. This result suggests that salivary anti-S1 and anti-NCP IgG testing with the same nLF platform could offer an effective and non-invasive approach to differentiate immune responses by vaccination (for mRNA vaccines targeting spike protein) or prior COVID-19 infections.

**Figure 3.**
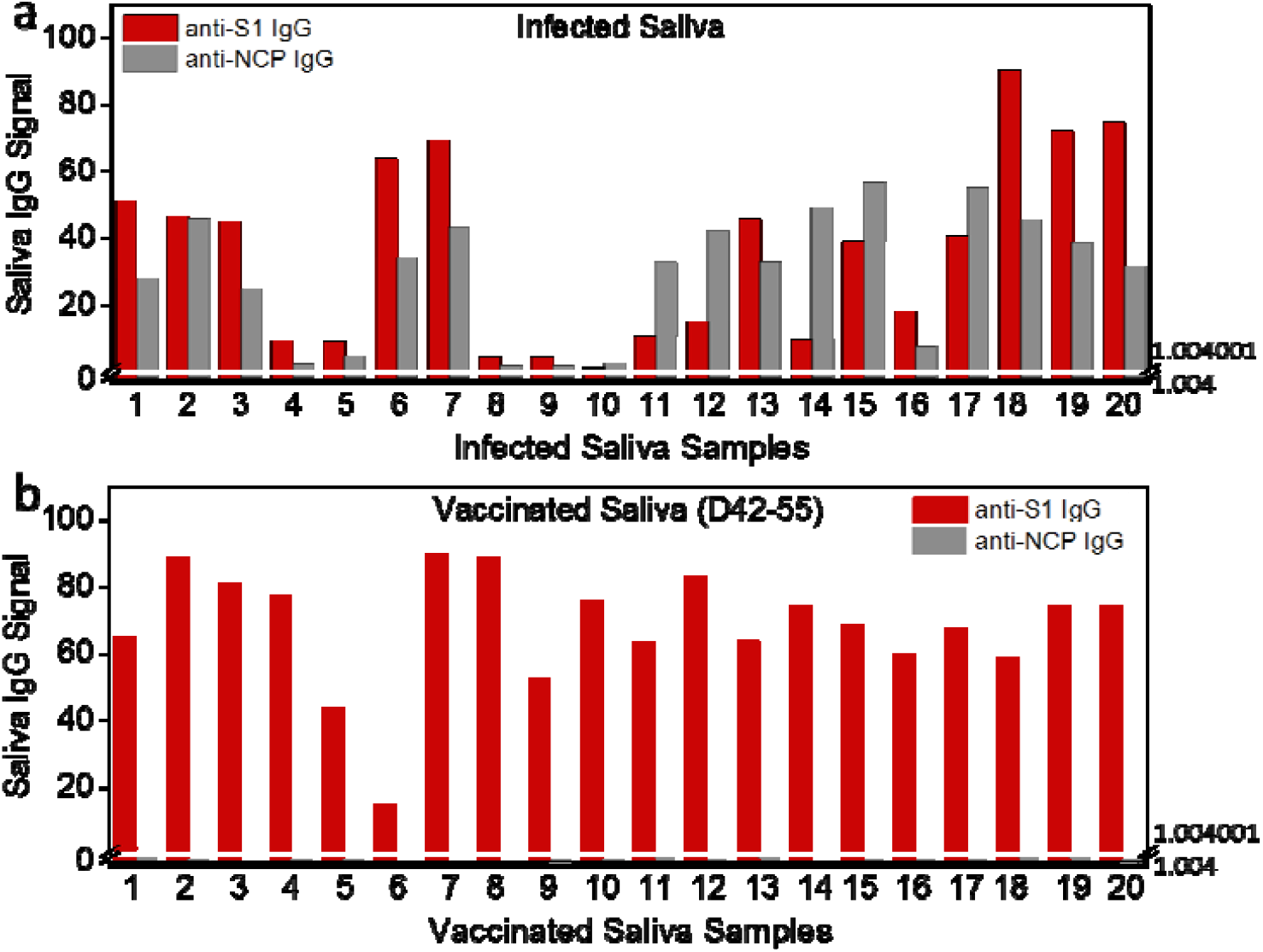
Salivary antibody patterns: vaccination vs. infection. **a**, Anti-S1 IgG and anti-NCP IgG signals for infected saliva samples. The cutoff value of 0.921 for anti-S1 IgG signal and 1.004 for anti-NCP IgG signal. All of the 20 COVID-19 infected saliva samples were tested positive for both anti-S1 IgG (100%) and anti-NCP IgG (100%). **b**, Anti-S1 IgG and anti-NCP IgG signals for vaccinated saliva samples. 20 vaccinated saliva samples were collected 42-55 days post 1st vaccine dose. Anti-S1 signal for all 20 vaccinated saliva samples are above the break line of 1.004 (> cutoff value of 0.921 for anti-S1 IgG signal) on Y-axis. Anti-NCP signal for all 20 vaccinated saliva samples are below the break line of 1.004 (= cutoff value for anti-NCP IgG signal) on Y-axis. Therefore, with 20 vaccinated saliva samples, 0 out of 20 (0%) is tested positive for anti-NCP IgG while all 20 (100%) are tested positive for anti-S1 IgG.

### Salivary antibody measured by nLF assay and antibodies in blood measured by a lab-based serology assay

To further validate our nLF assay for monitoring SARS-CoV-2 IgG trends post vaccination, we compared nLF IgG saliva test results with dried blood spot (DBS) serology test results using a previously established quantitative lab-test, the pGOLD assay ^11^. We longitudinally measured 64 previously COVID-19 free individuals who received mRNA vaccines (51 from Pfizer/BioNTech, 13 from Moderna) and donated matched saliva and DBS samples at multiple timepoints over ∼ 4 months. Samples were divided into 6 groups based on collection time after the 1st vaccine dose: D0 (before the 1st vaccine dose), D19-28 (19-28 days, right before the 2nd vaccine dose), and fully vaccinated groups D33-55 (33-55 days), D61-83 (61-83 days), D89-108 (89-108 days), and D112-133 (112-133 days). With nLF, salivary anti-S1 IgG positivity were 0% (0/20), 96.43% (27/28), 100% (30/30), 100% (49/49), 100% (14/14) and 100% (9/9) for the 6 groups respectively, corroborating with anti-S1 IgG positivity measured in matched DBS samples by the quantitative pGOLD assay **(Figure 4a)**. Only one vaccinated saliva sample from group D19-28 was not detected positive to anti-S1 IgG by the nLF assay, with its matched DBS sample also showing a much weaker signal than other samples in the same group by the pGOLD assay **(Figure S3)**. Follow-up studies of saliva and DBS samples from the same participant at the subsequent timepoint (D33-55) were tested anti-S1 IgG positive by nLF and pGOLD assays (data is not presented).

**Figure 4.**
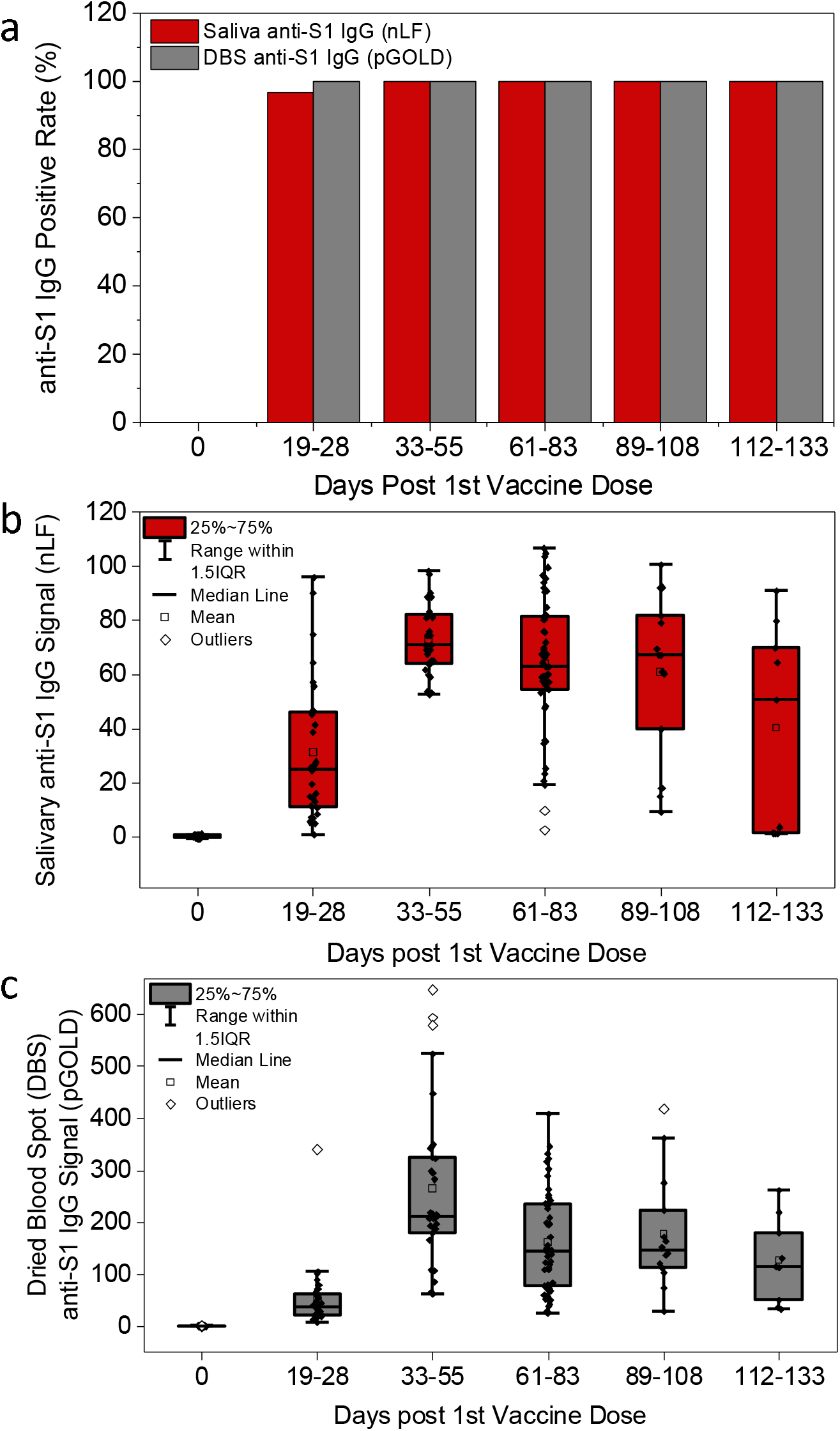
Comparison of salivary antibody measured by nLF assay and antibodies in blood measured by pGOLD assay. **a**, Anti-S1 IgG positive rate. Samples are divided into 6 groups: D0 (before the 1st vaccine dose), D19-28 (19-28 days, right before the 2nd vaccine dose), and fully vaccinated groups D33-55 (33-55 days), D61-83 (61-83 days), D89-108 (89-108 days), and D112-133 (112-133 days). With nLF, salivary anti-S1 IgG positivity are 0% (0/20), 96.43% (27/28), 100% (30/30), 100% (49/49), 100% (14/14) and 100% (9/9) for the 6 groups respectively. The anti-S1 IgG positivity measured in matched dried blood spot (DBS) samples by the pGOLD assay are 0% for D0 samples and 100% for all of the vaccinated groups. **b**, The evolution of salivary anti-S1 IgG monitored by nLF. **c**, The evolution of anti-S1 IgG levels in matched DBS samples monitored by pGOLD assay. Generally, anti-S1 IgG in both saliva and DBS showed moderate levels on day 19-28 after the first dose of mRNA vaccines, and further increased to higher levels after the second vaccine dose. The anti-S1 IgG levels reached a maximum on day 33-55, then remained elevated with a slow declining trend from the peak during the follow up timepoints (day 61-83, 89-108, and 112-133).

The evolution of salivary anti-S1 IgG and anti-S1 IgG levels in matched DBS samples were monitored by nLF and pGOLD respectively over ∼ 4 months post vaccination, showing similar trends **(Figure 4b**-**c)**. Generally, anti-S1 IgG in both saliva and DBS showed moderate levels on day 19-28 after the first dose of mRNA vaccines, and further increased to higher levels after the second vaccine dose. The anti-S1 IgG levels reached a maximum on day 33-55, then remained elevated with a slow declining trend from the peak during the follow up timepoints (day 61-83, 89-108, and 112-133). This trend was consistent with a recent report on antibody persistence over 6 months after the 2nd dose of the Moderna mRNA1273 vaccine ^39^. One disadvantage of the nLF assay, when compared to NIR fluorescence based pGOLD assay, is the narrower dynamic range of the test line intensity, making it less differentiating in the high end of antibody concentrations. Nevertheless, we observed a similar trend of anti-S1 IgG in saliva detected by nLF assay and anti-S1 IgG in DBS detected by pGOLD assay (**Figure 4**), suggesting the saliva-based assay using the nLF platform could represent a rapid, noninvasive alternative for SARS-CoV-2 IgG testing and longitudinal monitoring.

### Personalized monitoring of SARS-CoV-2 antibody persistence

To explore personalized monitoring of SARS-CoV-2 antibody persistence, we followed several vaccinated participants closely (5 from Pfizer/BioNTech, 2 from Moderna) by collecting their saliva samples weekly or bi-weekly and measuring their salivary anti-S1 IgG levels up to 13 weeks using our nLF assay. The antibody levels and details of evolution were highly variable among all participants **(Figure S4)**. Importantly however, the salivary and blood anti-S1 levels for individuals followed the same trend of evolution post vaccinations **(Figure S5)**, suggesting the feasibility of non-invasive salivary antibody measurements by nLF for longitudinal assessment of vaccination effects. For example, the anti-S1 IgG of participant A (Pfizer/BioNTech vaccine) was not detected until week 3. After the 2nd vaccine dose in week 3, the anti-S1 IgG declined in week 4 and then boosted in week 6 and 7, and declined later with some fluctuations **(Figure 5a)**. In the case of participant B, a moderate anti-S1 IgG was detected in week 2 post the 1st vaccine dose (Moderna), and the IgG level declined gradually in week 3 and 4. After the 2nd vaccine dose in week4, the IgG level jumped drastically in week 5, then was sustained at a relatively high level and sustained with modest decline from the peak during the following weeks **(Figure 5b)**.

**Figure 5.**
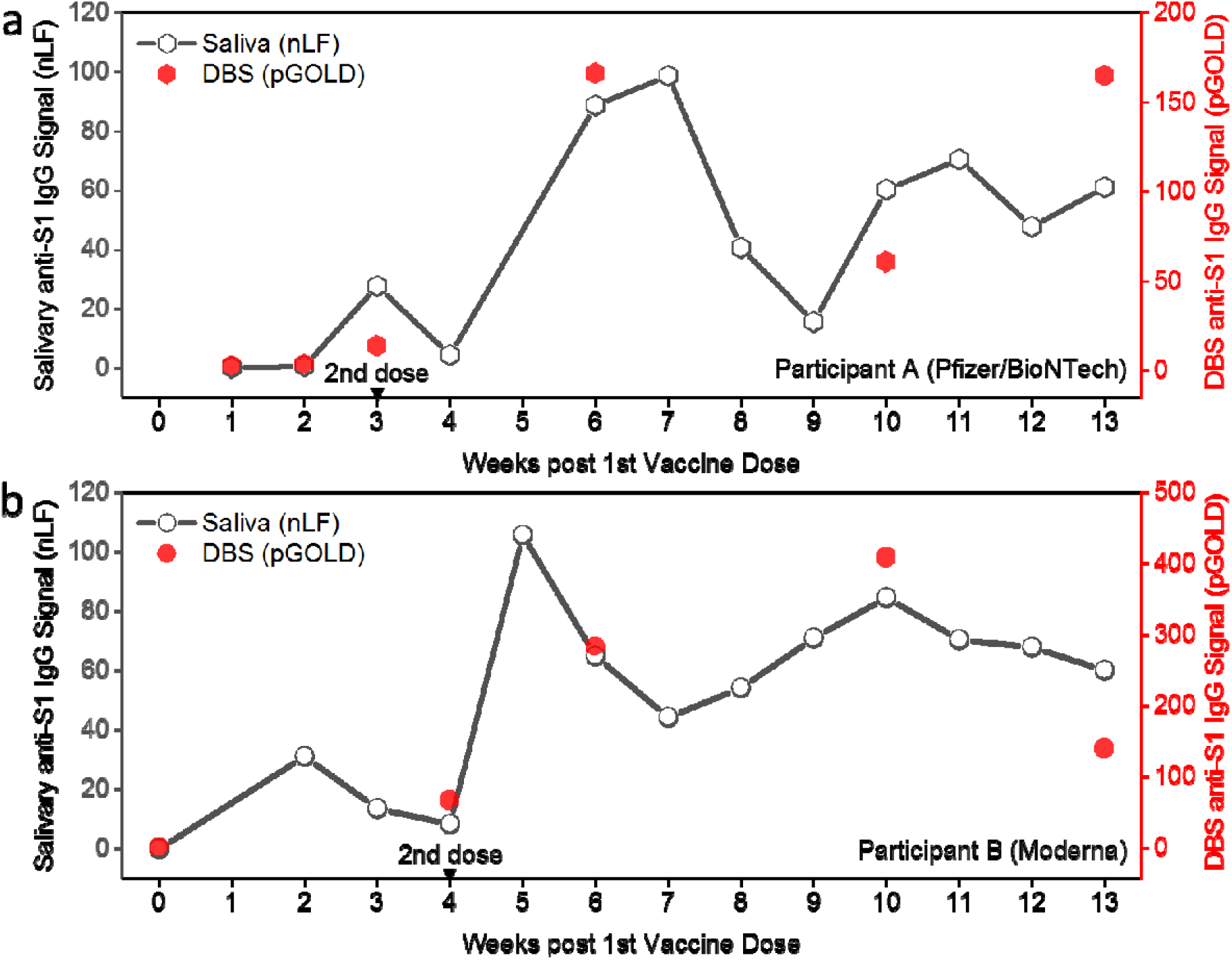
Personalized monitoring of SARS-CoV-2 antibody persistence. **a**, Monitoring of anti-S1 IgG over 13 weeks post vaccination of a representative participant who received Pfizer/BioNTech vaccine. Salivary anti-S1 IgG were collected and tested by nLF weekly or bi-weekly. Matched DBS samples were collected and tested by pGOLD assay at selected timepoints. **c**, Monitoring of anti-S1 IgG over 13 weeks post vaccination of a representative participant who received Moderna vaccine. Salivary anti-S1 IgG were collected and tested by nLF weekly or bi-weekly. Matched DBS samples were collected and tested by pGOLD assay at selected timepoints.

## Discussions

A simple but important feature of the nLF platform for rapid antibody detection is the removal of abundant, non-specific antibodies from a sample to afford higher analytical sensitivity (by ∼ 50-fold over conventional LF) and signal linearity. This added step can be done in POC settings using simple micro-centrifuging (handheld, battery operated one available) or magnetic separation by employing magnetic nanoparticles residing in the core of a gold shell and permanent magnets for rapid washing **(Figure S6)**. Although an additional step is required relative to conventional LF antibody test, it is crucial to obtain IgG signals on the test lines that reflect the specific antibody concentrations in the sample due to the elimination of competing antibodies that would always lower the true signal.

The nLF approach is applicable to detecting antibodies in all major body fluids, including serum, plasma, venous and capillary whole blood and dried blood spot, for assessing immune responses to a wide range of infectious diseases or vaccines in POC settings within 25 minutes. It is also useful for detecting autoantibodies for various autoimmune diseases. The rapid test can be semi-quantitative especially with the integration of an LF reader that detects light scattering of gold nanoparticles. A wide range of fluorescent or luminescent particles can also be employed for the nLF platform, boosting the nLF assay dynamic range by orders of magnitude for biomarker quantification.

We presented findings of longitudinal testing of salivary and blood IgG antibodies for individuals at various timepoints post vaccination up to ∼ 4 months. The study will continue for > 1 year to correlate salivary and blood antibody levels and their relations with immunity to COVID-19 infection. A limitation of our work is that rigorous immunity assessments require serological neutralizing antibody quantification, raising the question of relevance of anti-SARS-CoV-2 spike protein levels in saliva and blood. However, neutralization assays are time-consuming and employ live viruses, which are unsuitable for mass vaccination studies. Several simplified neutralization assays have been developed without using live viruses^20,40-42^. Importantly, there have been mounting evidence that neutralizing antibody levels in COVID-19 patients are well correlated with antibodies against spike protein on the surface of SARS-CoV-2^14,16,20-22^. Further, our results here showed that salivary anti-S1 IgG was well correlated with that in the blood for the mRNA vaccinated cohort as a whole (Figure 4) and at the individual level (Figure 5, Figure S4, and Figure S5). Such correlation has been seen recently for COVID-19 patients^23,24,27^ and in various other infectious diseases in the past few decades^30,31^. All of these results present a compelling case for monitoring salivary IgG against spike protein in vaccinated population as a non-invasive approach to assessing antibody levels and immunity. We clearly observed similar evolution trends and patterns for anti-S1 IgG in saliva and in blood. Since these antibodies are resulted entirely from vaccination, and when immunity wanes, salivary antibodies could also diminish. Continual monitoring of antibody persistence is crucial as COVID-19 vaccines circulate and aid the path to normalcy.

## Conclusions

Achieving > 99.5% sensitivity for anti-SARS-CoV-2 spike protein IgG detection in vaccinated saliva samples (19 days post first vaccine shot) at 100% specificity, the rapid nLF assay provides an efficient and easy tool to evaluate population immunity to COVID-19, identifying potential immunity gaps and susceptible populations to inform targeted vaccination/prevention strategies like a booster vaccine dose. The flexible designability of the nLF assay platform not only enables users to easily differentiate vaccination from infection, but also gives the potential to detect other infected and autoimmune diseases by adjusting the Au-detecting agent complexes. With further improvements and modifications of the nLF platform such as introducing magnetic particles and a LF test reader, the nLF assay will be more user friendly for POC and can be translated into home testing.

## Materials and Methods

### Biological samples and chemicals

The study was performed in accordance with standard ethical principles and approved by the LifeBridge Health local institutional review board under Protocol # 1707882 and clinical trials # 04910971. All patients provided written consent.

Saliva samples from 20 PCR-confirmed COVID-19 patients were collected at the Sinai Hospital of Baltimore. Paired saliva and dried blood spot (DBS) samples of 64 vaccinated participants (51 from Pfizer/BioNTech, 13 from Moderna) at multiple timepoints post vaccination were collected for the vaccine study. A 10 μl blood sample by fingerstick was collected and applied to DBS cards at various timepoints and stored at -20 °C before testing. At the same time of DBS sample collection, paired saliva samples were collected by a simple method of spitting into a plastic tube and stored at -20 °C before testing.

SARS-CoV-2 spike (S1) (40591-V08H) was purchased from Sino Biological. SARS-CoV-2 nucleocapsid protein (NCP) (ncov-PS-Ag6) was purchased from FAPON Biotech Inc.

All chemicals were purchased from Sigma-Aldrich, and were used as received without further purification, except where mentioned otherwise.

### Preparation of detection Au-antigen complex

A colloidal gold (Au) nanoparticle solution was prepared by adding 0.7 ml 1 wt.% sodium citrate solution to 80 ml boiling gold chloride solution (0.0125 wt.%). After a 30-minute reaction, the colloidal Au nanoparticles were prepared and cooled to room temperature for Au-antigen preparation. The magnetic Au (mAu) nanoparticle containing a Fe3O4 core and an Au shell are products of Nirmidas Biotech. Inc. For Au-antigen or mAu-antigen complex preparation, 10 μg of S1 or NCP was mixed with 1.25 ml colloidal Au or mAu solution for 30 minutes at room temperature, leading to the ionic adsorption of SARS-CoV-2 antigens on the surface of the colloidal Au or mAu particles. After blocking with a solution containing BSA, the mixture was centrifuged at 10000 rpm for 5 minutes. The supernatant was discarded and the pellet containing Au-antigen/mAu-antigen complex was suspended in 60 μL Tris buffer containing a surfactant and stored at -20 °C for later usage. With magnetic mAu particles, a magnetic rack was used to attract and hold the particle for removing unbound species and washing the particles with captured specific antibodies (see Figure S6).

### Fabrication of immunochromatographic strip

The immunochromatographic strip was composed of five components, a plastic backing, a sample pad, a conjugate pad, a nitrocellulose membrane and an absorbent pad. The sample pads and the conjugate pads were treated with 20 mM Tris buffer containing blocking agent and dried at 37°C and 25% relative humidity. The mouse anti human IgG (2 mg/ml) or the streptavidin (1 mg/ml) in PBS (1% wt. trehalose) was dispensed at the test or the control line on the nitrocellulose membrane, using a dispenser at a rate of 1.0 μL/cm and then dried at 37°C. The gold nanoparticles conjugated with biotin-BSA were applied to the treated conjugate pad and then dried completely. The absorption pad, nitrocellulose membrane, pretreated conjugate pad, and sample pad were attached to a plastic backing and assembled as a strip with a 1.5 mm overlap, sequentially. The assembled plate was cut into 4-mm-wide pieces, using an automatic strip cutter ZQ2000. The generated strip products were packaged in a plastic bag with desiccant and stored at room temperature.

### nLF assay for SARS-CoV-2 IgG antibody detection

To test a saliva sample, 200 μl of saliva was mixed with 10 μl Au-S1 and 50 μl Tris buffer in a 1.5 ml microcentrifuge tube. To test a serum sample, 5 μl serum was mixed with 10 μl Au-S1 and 390 μl Tris buffer in a 1.5 ml microcentrifuge tube. After a 10-minute incubation at room temperature, 1 ml Tris buffer was added to the tube and the mixture was centrifuged in a mini centrifuge for 5 minutes. The supernatant was discarded, and the pellet was suspended in 100 μL PBS running buffer to add to the sample well in the assay card. The result was read and photographed after 5 minutes of migration of the added sample through the test strip. The test line image intensity and background intensity were read and analyzed by ImageJ. The test line signal level of nLF was determined by the test line image intensity subtracted by background intensity.

### pGOLD™ SARS-CoV-2 IgG antibody assay

A 2 mm circle was punched out from a DBS sample (comprised of 10 μl dried finger stick whole blood) and dissolved in 160 µL sample buffer for 1 h. After centrifugation at 3000 g for 5 minUTES, the DBS-dissolved sample solution was heat inactivated at 56 °C for 30 minutes. The pGOLD antibody assay was performed in a 64-well format following a protocol similar to our previous work^11^.

## Data Availability

All data referred to in the manuscript are available upon request to the corresponding author.

## ACKNOWLEDGMENTS

This work was supported by Platelet and Thrombosis Research, LLC, Baltimore, MD, USA. Device fabrication was supported by Shenzhen Grant No. JSGG20191231141403880 and Guangdong Grant No. 2020B1111160003.

## Supplementary information

**Figure S1.**
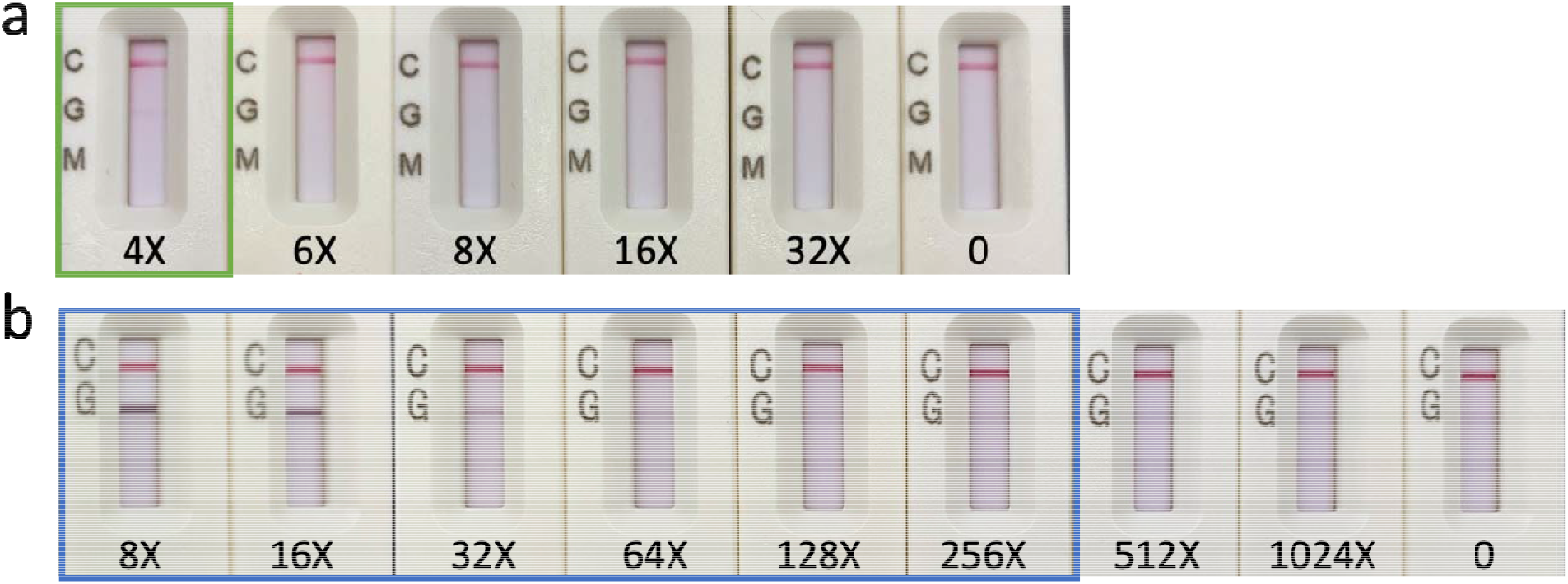
Detection limit of anti-spike IgG in serum by nLF and MidaSpot. Purified recombinant anti-SARS-CoV-2 spike protein IgG was spiked into a negative serum sample and conducted serial dilutions (4X to 1024X dilutions) to determine the detection limit by nLF and MidaSpot. **a**, Detection limit of anti-spike IgG in serum by MidaSpot. Nirmidas’ MidaSpot™ COVID-19 rapid Antibody Combo Detection Kit was approved by FDA EUA for POC testing. Based on the testing instruction, 10 µl serum sample was applied to the sample window followed by adding 4 drops (∼ 100 µl) of running buffer, and after 20 min the result was recorded. MidaSpot presents positive signals for the 4X and lower dilutions. **b**, Detection limit of anti-spike IgG in serum by nLF. The nLF (see Method) is able to detect anti-spike IgG even in the 256x diluted serum sample, affording a ∼ 64 times lower detection limit than the conventional LF approach owing to the removal of excess, non-specific serum IgG by nLF.

**Figure S2.**
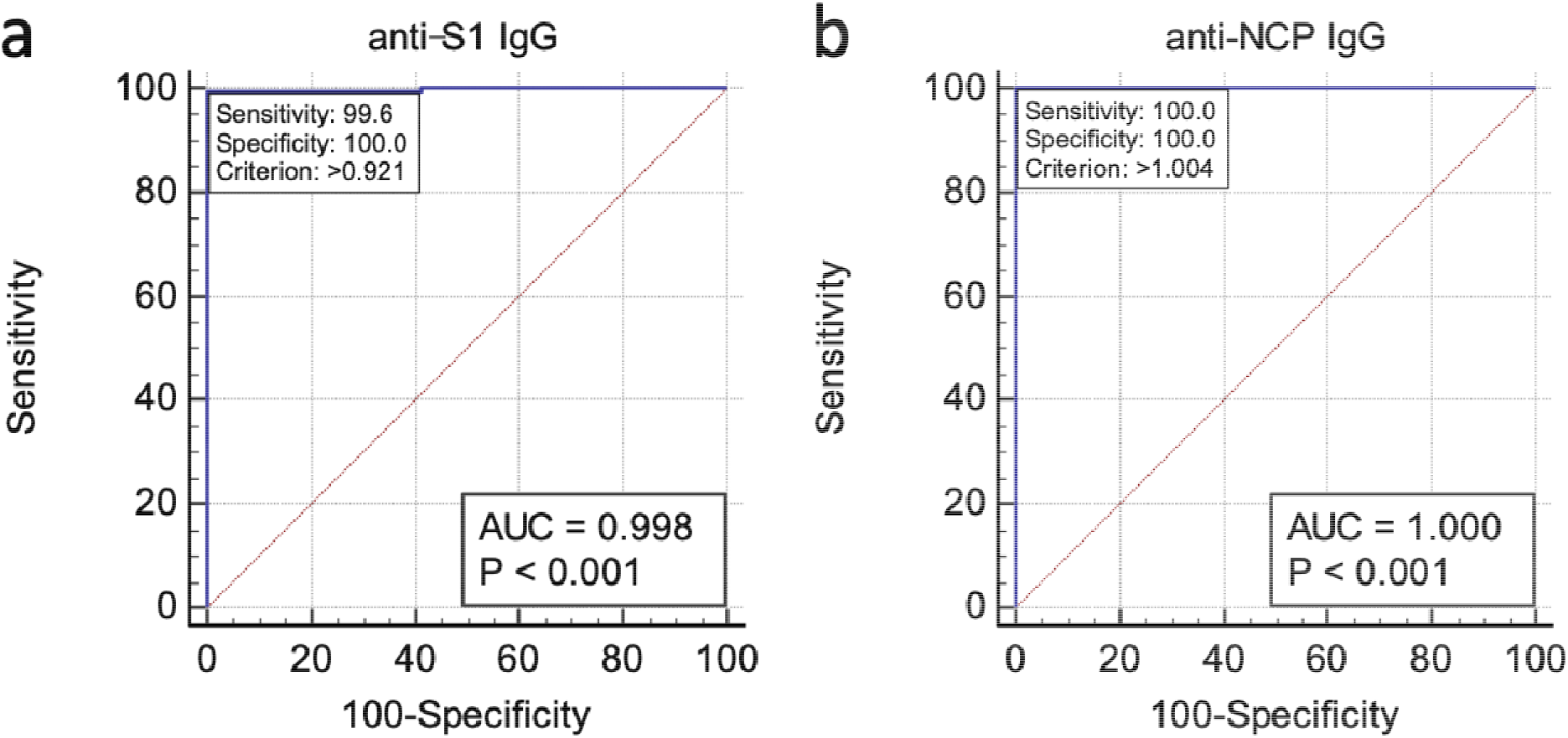
ROC curves for the nLF SARS-CoV-2 IgG assay. **a**, ROC curve for the nLF SARS-CoV-2 anti-S1 IgG assay. The ROC curve is based on 34 unvaccinated healthy saliva samples without prior COVID-19 infection, 20 saliva samples from PCR confirmed COVID-19 patients (16 samples were collected ≥ 15 days post symptom onset, 1 sample was collected 11 days post symptom onset, and 3 samples had no information regarding the number of days between symptom onset to sample collection), and 205 vaccinated (with mRNA vaccines from Pfizer/BioNTech and Moderna) saliva samples (collected ≥ 19 days post 1st vaccine shot from subjects without prior COVID-19 infection). A cut-off value of 0.921 T-line intensity unit was determined via the ROC curve under the criteria of 100% specificity and 99.6% sensitivity. **b**, ROC curve for the nLF SARS-CoV-2 anti-NCP IgG assay. The ROC curve is based on 12 negative and 20 PCR-positive COVID-19 saliva samples. The ROC analysis gave a cutoff value of 1.004 intensity unit under the criteria of 100% specificity and 100% sensitivity.

**Figure S3.**
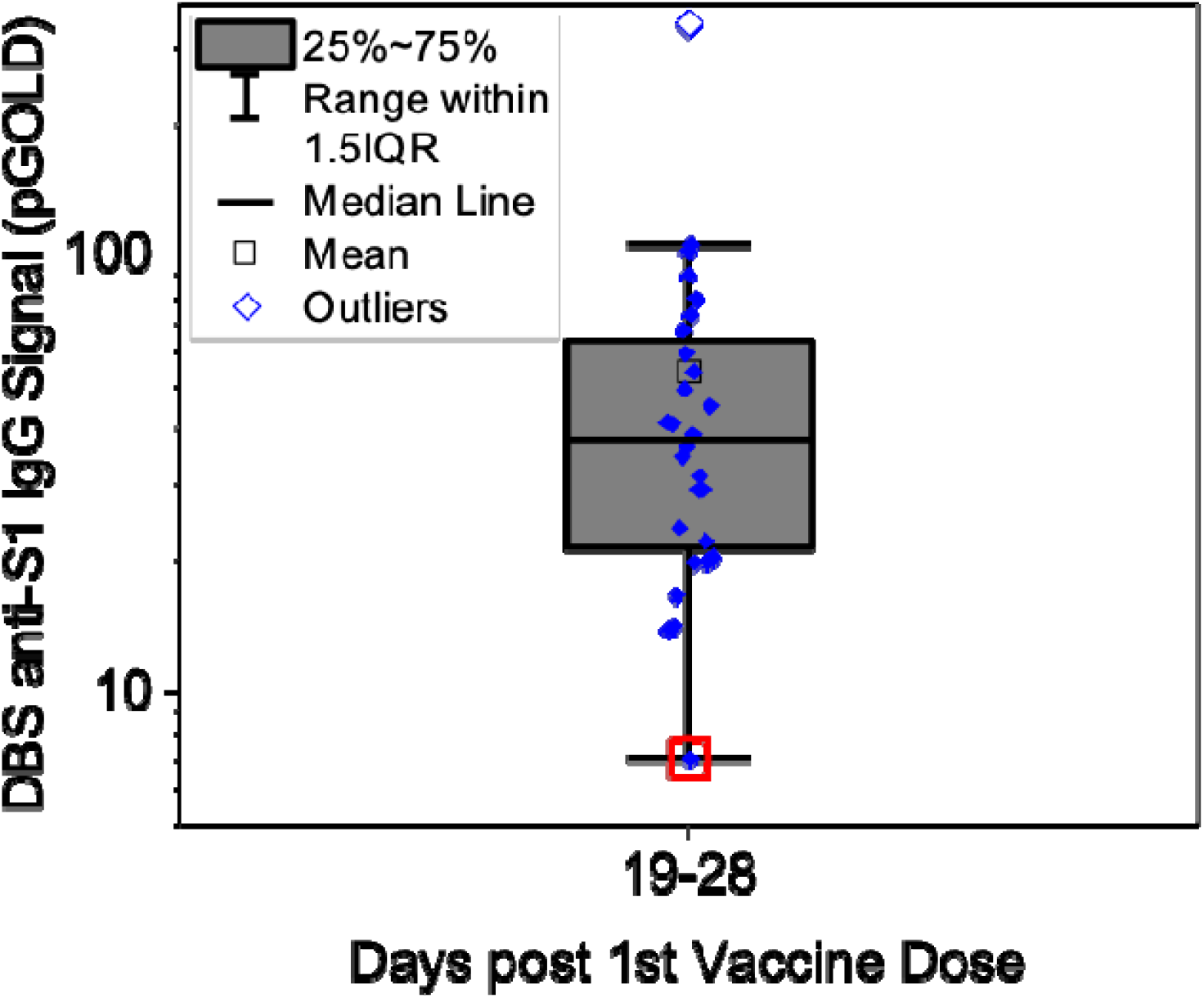
pGOLD results for anti-S1 IgG of DBS samples from group D19-28. 28 saliva samples from the group D19-28 are tested by nLF, and their matched DBS samples are tested by pGOLD assay. 1 out of 28 vaccinated saliva samples was not detected positive to anti-S1 IgG by the nLF, with its matched DBS sample (red square) also showing a much weaker signal than other samples in the same group by the pGOLD assay.

**Figure S4.**
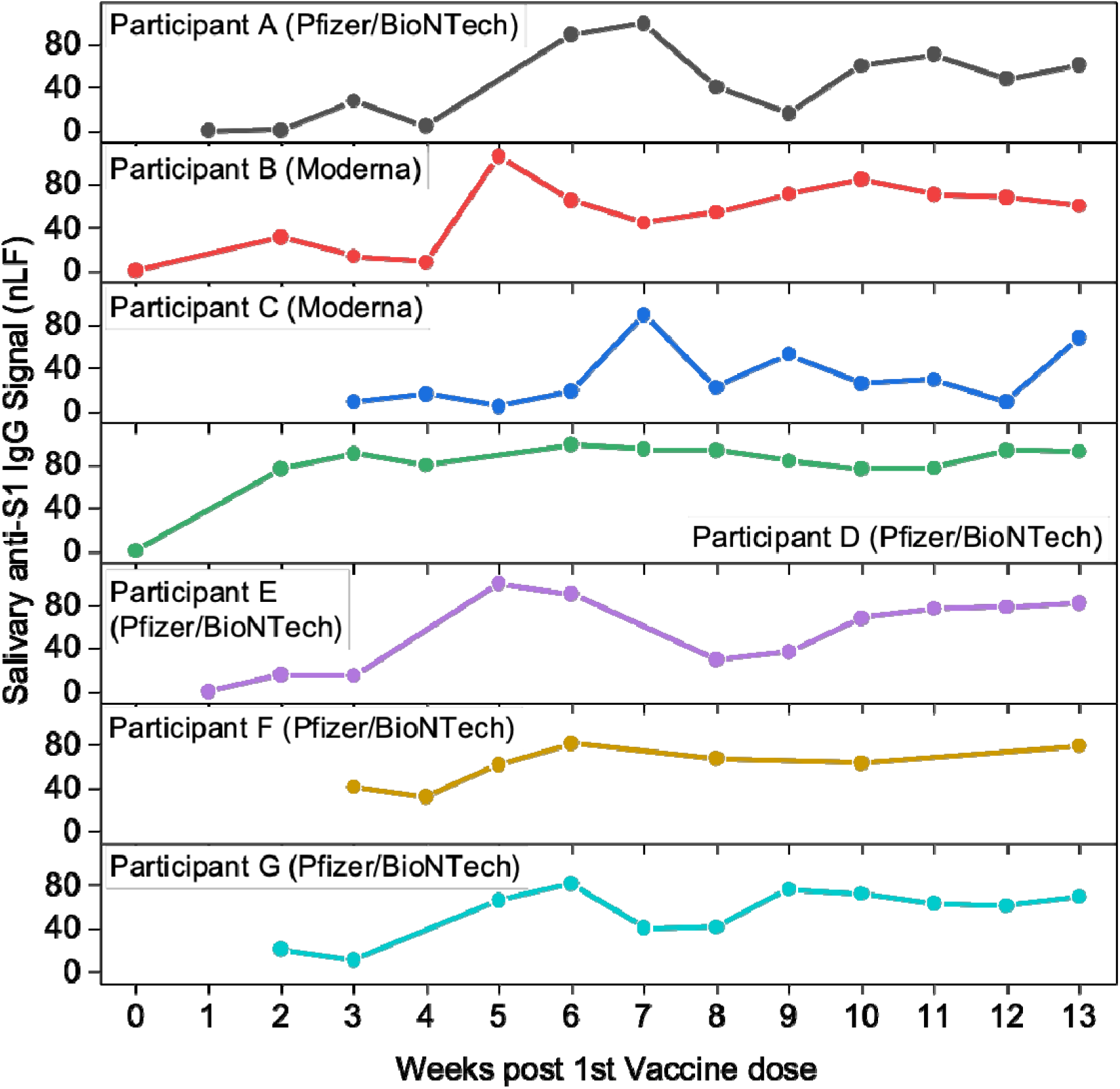
Personalized monitoring of SARS-CoV-2 antibody persistence. Personalized monitoring of SARS-CoV-2 anti-S1 IgG levels for 7 vaccinated participants (participant A-G) (5 from Pfizer/BioNTech, 2 from Moderna) up to 13 weeks using the nLF assay.

**Figure S5.**
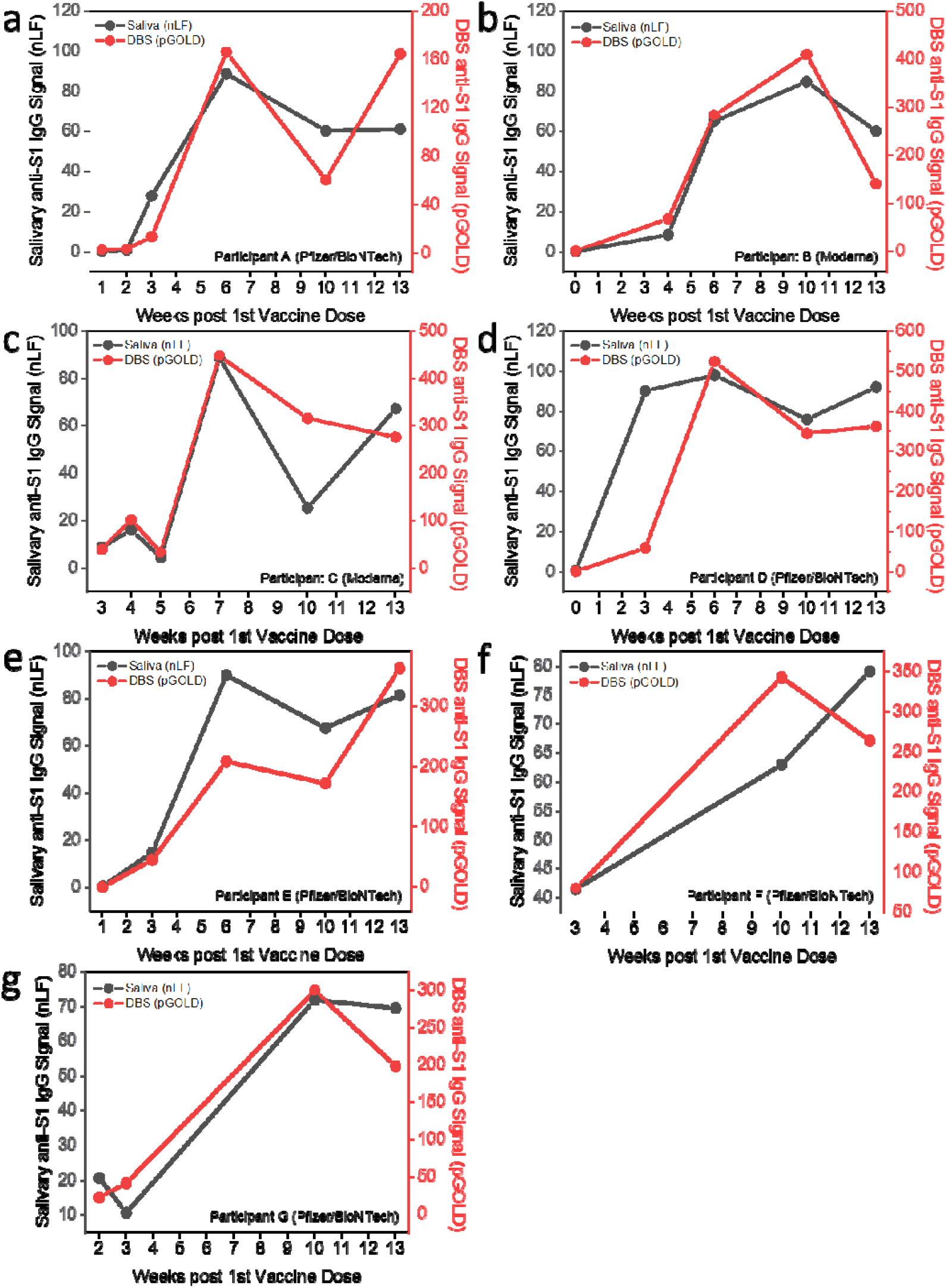
Comparison of salivary and blood anti-S1 IgG post vaccination for individuals. **a-g**, Comparison of salivary and blood anti-S1 IgG for 7 individuals (5 from Pfizer/BioNTech, 2 from Moderna) at selected timepoints post vaccination. Salivary anti-S1 IgG signals were detected by nLF. DBS anti-S1 IgG signals were detected by pGOLD assay using DBS samples.

**Figure S6.**
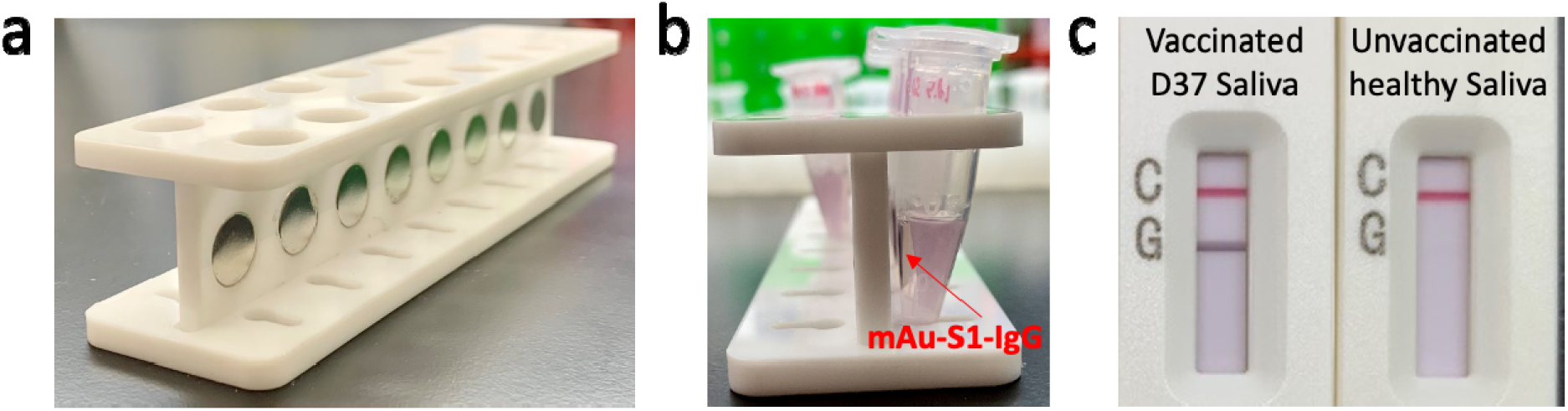
Magnetic Au nanoparticle (mAu) based nLF antibody assay. **a**, A magnetic rack for removing unbound non-specific antibodies. **b**, mAu-S1-IgG complex collection by the magnetic rack. **c**, Representative results from mAu based nLF for anti-S1 IgG positive and negative saliva samples.

## References

1. Ravens-Sieberer, U., et al. Impact of the COVID-19 pandemic on quality of life and mental health in children and adolescents in Germany. Eur Child Adolesc Psychiatry (2021).

2. Fontanet, A., et al. SARS-CoV-2 variants and ending the COVID-19 pandemic. The Lancet 397, 952–954 (2021).

3. Salyer, S.J., et al. The first and second waves of the COVID-19 pandemic in Africa: a crosssectional study. The Lancet 397, 1265–1275 (2021).

4. Padhan, R. & Prabheesh, K.P. The economics of COVID-19 pandemic: A survey. Econ Anal Policy 70, 220–237 (2021).

5. Skegg, D., et al. Future scenarios for the COVID-19 pandemic. The Lancet 397, 777–778 (2021).

6. Tanaka, T. & Okamoto, S. Increase in suicide following an initial decline during the COVID-19 pandemic in Japan. Nat Hum Behav 5, 229–238 (2021).

7. Loomba, S., de Figueiredo, A., Piatek, S.J., de Graaf, K. & Larson, H.J. Measuring the impact of COVID-19 vaccine misinformation on vaccination intent in the UK and USA. Nat Hum Behav 5, 337–348 (2021).

8. Keehner, J., et al. SARS-CoV-2 infection after vaccination in health care workers in California. N Engl J Med 384, 1774–1775 (2021).

9. Edward P. K. Parker, M.S., Beate Kampmann. Keeping track of the SARS-CoV-2 vaccine pipeline. Nat Rev Immunol 20, 1 (2020).

10. Van Elslande, J., et al. Diagnostic performance of seven rapid IgG/IgM antibody tests and the Euroimmun IgA/IgG ELISA in COVID-19 patients. Clin Microbiol Infect 26, 1082–1087 (2020).

11. Liu, T., et al. Quantification of antibody avidities and accurate detection of SARS-CoV-2 antibodies in serum and saliva on plasmonic substrates. Nat Biomed Eng 4, 1188–1196 (2020).

12. Cai, X.F., et al. A peptide-based magnetic chemiluminescence enzyme immunoassay for serological diagnosis of coronavirus Disease 2019. J Infect Dis 222, 189–193 (2020).

13. Wu, J.L., et al. Four point-of-care lateral flow immunoassays for diagnosis of COVID-19 and for assessing dynamics of antibody responses to SARS-CoV-2. J Infect 81, 435–442 (2020).

14. Wajnberg, A., et al. Robust neutralizing antibodies to SARS-CoV-2 infection persist for months. Science 370, 1227–1230 (2020).

15. To, K.K.-W., et al. Temporal profiles of viral load in posterior oropharyngeal saliva samples and serum antibody responses during infection by SARS-CoV-2: an observational cohort study. The Lancet Infectious Diseases 20, 565–574 (2020).

16. Yang, Y. & Du, L. SARS-CoV-2 spike protein: a key target for eliciting persistent neutralizing antibodies. Signal Transduct Target Ther 6, 95 (2021).

17. Wang, N., Shang, J., Jiang, S. & Du, L. Subunit Vaccines Against Emerging Pathogenic Human Coronaviruses. Front Microbiol 11, 298 (2020).

18. Salazar, E., et al. Convalescent plasma anti-SARS-CoV-2 spike protein ectodomain and receptor-binding domain IgG correlate with virus neutralization. J Clin Invest 130, 6728–6738 (2020).

19. Iyer, A.S., et al. Persistence and decay of human antibody responses to the receptor binding domain of SARS-CoV-2 spike protein in COVID-19 patients. Sci Immunol 5(2020).

20. Nie, J., et al. Quantification of SARS-CoV-2 neutralizing antibody by a pseudotyped virus-based assay. Nat Protoc 15, 3699–3715 (2020).

21. Tan, C.W., et al. A SARS-CoV-2 surrogate virus neutralization test based on antibody-mediated blockage of ACE2-spike protein-protein interaction. Nature biotechnology 38, 1073–1078 (2020).

22. Byrnes, J.R., et al. Competitive SARS-CoV-2 Serology Reveals Most Antibodies Targeting the Spike Receptor-Binding Domain Compete for ACE2 Binding. mSphere 5(2020).

23. MacMullan, M.A., et al. ELISA detection of SARS-CoV-2 antibodies in saliva. Sci Rep 10, 20818 (2020).

24. Pisanic, N., et al. COVID-19 serology at population scale: SARS-CoV-2-specific antibody responses in saliva. J Clin Microbiol 59(2020).

25. Ter-Ovanesyan, D., et al. Ultrasensitive measurement of both SARS-CoV-2 RNA and antibodies from saliva. Anal Chem 93, 5365–5370 (2021).

26. Hettegger, P., et al. High similarity of IgG antibody profiles in blood and saliva opens opportunities for saliva based serology. PLoS One 14, e0218456 (2019).

27. Isho, B., et al. Persistence of serum and saliva antibody responses to SARS-CoV-2 spike antigens in COVID-19 patients. Sci Immunol 5(2020).

28. Isho, B., et al. Evidence for sustained mucosal and systemic antibody responses to SARSCoV-2 antigens in COVID-19 patients. medRxiv (2020).

29. Chakravarti, A., Matlani, M. & Jain, M. Immunodiagnosis of dengue virus infection using saliva. Curr Microbiol 55, 461–464 (2007).

30. Li, X., et al. Plasmonic gold chips for the diagnosis of Toxoplasma gondii, CMV, and rubella infections using saliva with serum detection precision. Eur J Clin Microbiol Infect Dis 38, 883-890 (2019).

31. Yoshizawa, J.M., et al. Salivary biomarkers: toward future clinical and diagnostic utilities. Clin Microbiol Rev 26, 781–791 (2013).

32. Michailidou, E., Poulopoulos, A. & Tzimagiorgis, G. Salivary diagnostics of the novel coronavirus SARS-CoV-2 (COVID-19). Oral Dis (2020).

33. Ketas, T.J., et al. Antibody responses to SARS-CoV-2 mRNA vaccines are detectable in saliva. bioRxiv (2021).

34. Koczula, K.M. & Gallotta, A. Lateral flow assays. Essays Biochem 60, 111–120 (2016).

35. Ebinger, J.E., et al. Antibody responses to the BNT162b2 mRNA vaccine in individuals previously infected with SARS-CoV-2. Nat Med (2021).

36. Sahin, U., et al. COVID-19 vaccine BNT162b1 elicits human antibody and TH1 T cell responses. Nature 586, 594–599 (2020).

37. Wang, Z., et al. mRNA vaccine-elicited antibodies to SARS-CoV-2 and circulating variants. Nature 592, 616–622 (2021).

38. Strömer, A., et al. Kinetics of nucleo- and spike protein-specific immunoglobulin G and of virus-neutralizing antibodies after SARS-CoV-2 infection. Microorganisms 8(2020).

39. Doria⍰Zrose, N., et al. Antibody persistence through 6 months after the second dose of mRNA-1273 vaccine for Covid-19. N Engl J Med, 3 (2021).

40. Cantoni, D., Mayora-Neto, M. & Temperton, N. The role of pseudotype neutralization assays in understanding SARS CoV-2. Oxf Open Immunol 2, iqab005 (2021).

41. Tani, H., et al. Evaluation of SARS-CoV-2 neutralizing antibodies using a vesicular stomatitis virus possessing SARS-CoV-2 spike protein. Virol J 18, 16 (2021).

42. Zheng, Y., et al. Neutralization assay with SARS-CoV-1 and SARS-CoV-2 spike pseudotyped murine leukemia virions. Virol J 18, 1 (2021).

